# Wild-type *C9orf72* expression is a genetic modifier of C9-ALS survival

**DOI:** 10.64898/2026.02.06.26345684

**Authors:** Stanislav Tsitkov, Akshay Raju, Jie Wu, Jonathan Li, Ryan G Lim, Zhuoxing Wu, Noura Al Bistami, Answer ALS Consortium, Jennifer E. Van Eyk, Clive N Svendsen, Jeffrey D Rothstein, Jonathan D Glass, Steve Finkbeiner, Julia A Kaye, Leslie M Thompson, Ernest Fraenkel

## Abstract

Amyotrophic lateral sclerosis (ALS) is highly heritable, yet the vast majority of cases lack an identifiable genetic cause and clinical progression remains largely unpredictable. To connect noncoding and rare genetic variation to disease phenotypes in a relevant cell type, we generated a multi-omic quantitative trait locus (QTL) atlas from 594 induced-pluripotent-stem-cell–derived human motor neuron lines (522 ALS patients, 72 controls). By mapping cis-QTLs for chromatin accessibility, splicing and gene expression from whole-genome sequencing, we identify common and rare variants on the wild-type *C9orf72* allele that form regulatory haplotypes. These haplotypes influence *C9orf72* expression levels in motor neurons and stratify C9-ALS patients into four subgroups; using clinical disease duration data and longitudinal ALSFRS-R scores, we show that these subgroups exhibit different survival trajectories, indicating that wild-type *C9orf72* expression acts as a genetic modifier of disease duration. Beyond the *C9orf72* locus, we detect ultra-rare intronic variants that create cryptic exons and structural and nonsense variants in established ALS genes, providing likely genetic explanations for disease in additional patients who previously lacked a molecular diagnosis. Our results show that QTL mapping in patient-derived motor neurons can reveal regulatory modifiers of progression and hidden pathogenic events in ALS, providing a framework for genetically informed risk attribution and patient stratification in complex neurological diseases.

## 2 Introduction

Amyotrophic lateral sclerosis (ALS) is a neurodegenerative disorder characterized by progressive motor impairments and motor neuron loss^1^. Approximately 15% of all cases are inherited, and causal genomic mutations in over two dozen genes have been identified^2^, yet even among genetically defined subtypes, rates of clinical progression vary widely and remain poorly explained. As most of these mutations have been discovered using family-based linkage studies and exome sequencing, almost all are protein coding^3^. However, there is likely additional genomic variation located in the non-protein coding regions that can contribute to risk of disease and phenotypic heterogeneity as well; these variants can disrupt DNA- and RNA-protein interactions, leading to dysfunction in transcription, splicing, or translation. With the expanding availability of genomics data from ALS patients and the use of genome wide association studies (GWAS)^4^, there is now potential to identify non-coding mutations causing disease. For example, two common intronic variants in *UNC13A*, (among the top GWAS hits^4^), were found to increase patient vulnerability by altering cryptic exonic splicing^5^.

Despite these advances, elucidating the functional impact of non-coding variants remains challenging, particularly for ultra-rare variants and for those whose effects may manifest as quantitative differences in disease course rather than binary disease risk. Regulatory regions are comparatively understudied, non-coding variants often exert small and context-dependent effects that are difficult to detect and validate experimentally, and many regulatory elements are under weak evolutionary constraint, making it difficult to distinguish true functional signals from background genomic variation using genomic data alone.

In this study, we overcome this challenge by mapping genomic variation to measured molecular phenotypes using Quantitative Trait Loci (QTL) analyses from data generated from Answer ALS (AALS)^6^. Among known ALS genes, C9orf72 is a natural focus because the hexanucleotide repeat expansion is the most common known genetic cause of ALS. As a result, C9-ALS has been the subject of multiple clinical trials^7^, underscoring both the need to understand sources of clinical heterogeneity within this population and the importance of genetically informed patient stratification. The size of the Answer ALS dataset and the broad inclusion criteria also provide an opportunity to search for rare, previously unrecognized genetic causes of ALS.

As part of AALS we generated RNA-seq and ATAC-seq profiles of induced pluripotent stem cell (iPSC)-derived motor neurons (diMNs) and matched whole-genome sequencing (WGS), expanding our prior dataset^6,8,9^ to a total of 594 individuals. Unlike post-mortem tissue, which tends to reflect late stages of disease, iPSCs allow us to model relationships between genomics and gene regulation and expression that may reflect early, causal events in the etiology of ALS. We use these sequencing data to generate tables of QTLs: genomic variants whose genotype is significantly associated with variation in gene expression (eQTLs), gene splicing (sQTLs), or chromatin accessibility (caQTLs).

We used our QTLs to explore potential functional consequences of risk variants identified in Project MinE’s ALS GWAS study^4^. First, we identified a linkage-disequilibrium (LD) block of significant Project MinE GWAS variants that were identified as expression QTLs (eQTLs) for *SCFD1* and splicing QTLs (sQTLs) for exon-skipping of *G2E3* exon 2. Leveraging transcription factor footprinting analysis using the ATAC-seq data, we identified the rs229143 variant as the probable causal *SCFD1* eQTL, likely functioning by disrupting DNA binding by the transcription factor ISL1 in a region of accessible chromatin in *SCFD1* intron 1.

We also used our QTLs to gain insight into known ALS subtypes. First, we found that there were several highly significant eQTLs for *C9orf72* expression. Combining these eQTLs with a genomic haplotype analysis at the *C9orf72* locus, we found that C9-ALS patients could be stratified into four groups characterized by the eQTL genotype of the wild-type allele that does not carry the mutant repeat expansion. Extending our analysis with the ALS Compute database^10–14^, we show that these groups are significantly associated with disease progression rates, as measured both by disease duration and the ALSFRS-R Total scale. Altogether, we show that genotype-driven upregulation of the *C9orf72* wild-type allele is significantly associated with slower rates of disease progression.

Finally, by examining extreme expression variation in genes known to harbor genetic causes of ALS, we discovered a number of genomic variants that may be patient-specific causes of disease: ultrarare intronic SNPs that induce cryptic-exon retention and create loss-of-function alleles in *TBK1* and *SPG11*, while structural deletions and nonsense mutations converge to silence *NEK1*, *OPTN* and other established ALS genes.

Our analysis demonstrates how the integration of genomic, epigenomic, and transcriptomic data gathered from diMNs from ALS patients and healthy controls can reveal previously unrecognized causes of ALS and help unravel likely trajectories of disease, which would help patients understand their disease course, and ultimately pave the way for novel targeted therapeutic strategies.

## 3 Results

### 3.1 Genomic, epigenomic, and transcriptomic data were collected for 594 iPSC-derived motor neuron lines

The primary goal of this study is to better understand the complex genetic underpinnings of ALS. Direct biopsy of motor neurons from living patients is not possible as it requires subjecting patients to risky procedures without a clear clinical benefit, and post-mortem tissue reflects end-stage pathology, limiting insight into earlier disease mechanisms. To model early ALS, we used diMNs (Figure 1a), which provide a physiologically relevant model that can be systematically obtained from living individuals.

**Figure 1:**
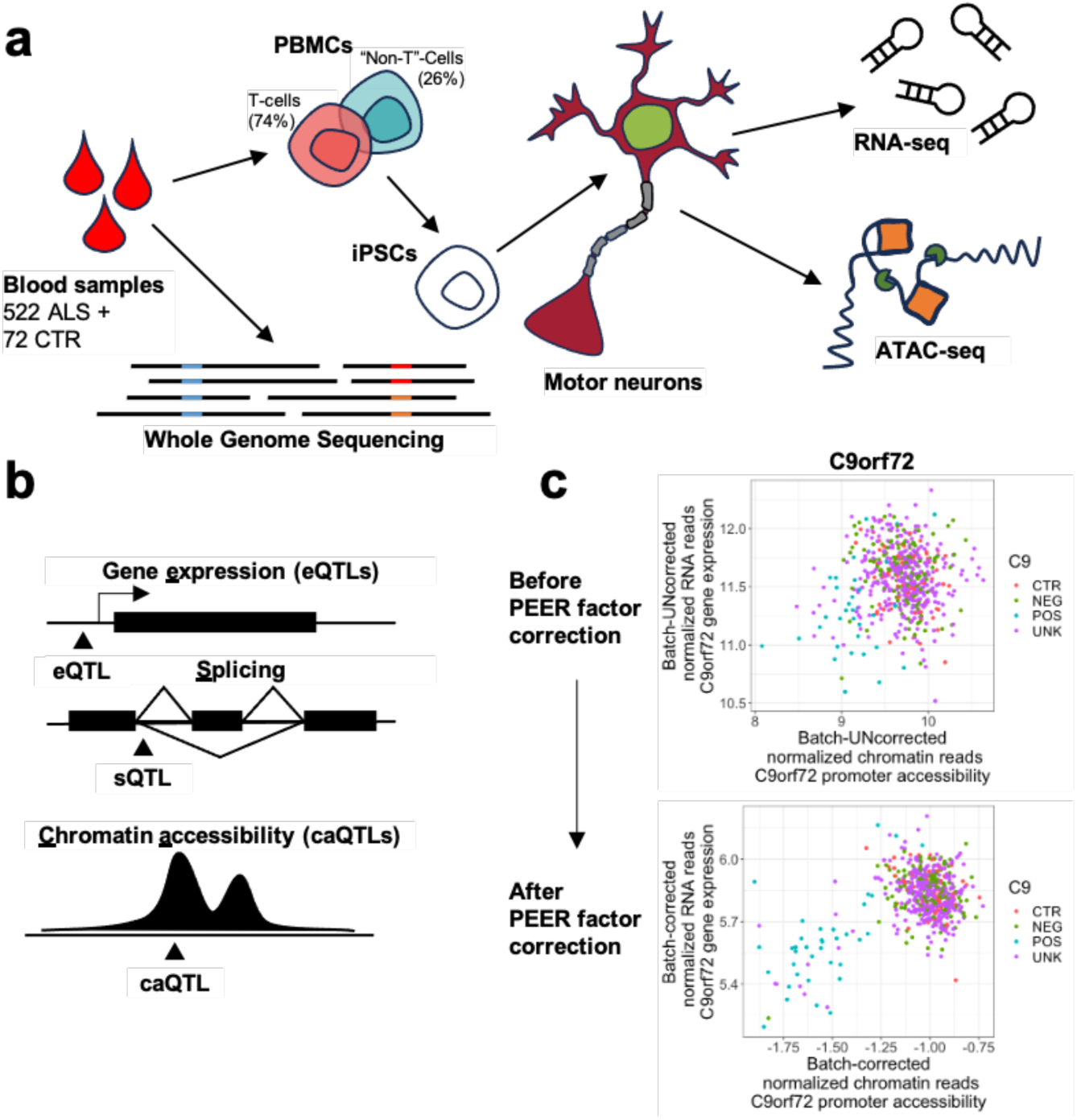
Overview of study design and batch correction. A) Schematic depicting the generation of ATAC-seq and RNA-seq data by AALS. WGS was performed on blood derived from ALS patients and healthy controls. PBMCs from blood are reprogrammed into iPSCs, which are in turn differentiated into diMNs. diMN cultures are then subjected to ATAC-seq and RNA-seq. B) Cartoon depicting QTLs generated in this study. eQTLs correspond to genomic variants that affect gene expression, sQTLs correspond to genomic variants that affect splicing, and caQTLs correspond to genomic variants that affect chromatin accessibility. C) The C9orf72 gene exhibits an increase in the significance of the association between expression and promoter accessibility after regressing out PEER factors; interestingly, over 80% of all C9-ALS cases separate out from controls and other ALS cases after correction. Point colors correspond to information in clinical metadata regarding presence of mutant repeat expansion in that sample: ALS samples carrying mutant repeat expansion (POS), ALS samples not carrying mutant repeat expansion (NEG), unlabeled ALS samples (UNK), and healthy controls (CTR).

diMNs were generated from iPSCs reprogrammed from blood samples of 594 individuals (522 ALS, 72 controls) and extensively profiled by AALS (Figure 1a); a 460-sample subset of these lines has been examined previously^6,8,9^. WGS was performed on whole blood from each individual, and epigenomic (ATAC-seq) and transcriptomic (RNA-seq) data were collected from each diMN line (Figure 1a). ATAC-seq data were used to identify open chromatin regions (peaks) and quantify chromatin accessibility to protein binding. A consensus peakset consisting of 101,430 chromatin regions was assembled by retaining open chromatin regions that were accessible in at least 10% of all samples. RNA-seq data was used to quantify gene expression and intron usage frequency. There were 26,971 transcripts remaining after filtering out those that were poorly expressed. We also measured 125,816 introns which corresponded to 26,606 AS events (Methods). The genotypes of 6,416 genomic variants falling both in exons and peaks of high read coverage were called and compared to WGS to verify sample concordance.

The cohort was roughly evenly split by sex (339 male, 255 female, Figure S1a-b), mostly of European ancestry (Figure S1c), and iPSCs for most lines were derived from T-cells (Figure S1a-b). Clinical metadata gathered on AALS participants indicated that out of the 522 individuals with ALS, 36 were carriers of the mutant *C9orf72* hexanucleotide repeat expansion (HRE) and 12 were carriers of deleterious mutations in *SOD1 (*Table S1). Only 63 reported a family history of the disease (Figure S1d-e). As the family history records were incomplete, the actual number may be slightly higher (see methods).

### 3.2 Accounting for hidden batch effects maximizes numbers of detected QTLs and identifies subgroups of ALS patients associated with C9orf72

Genome-driven variation in chromatin accessibility and transcription may be masked by alternative sources of variation, including batch effects, patient demographics, cell-type composition, and biological regulation. Our previous analyses of the AALS data have revealed a number of covariates that are significantly associated with both diMN gene expression and chromatin accessibility^8,9^. Further yet uncharacterized, hidden, batch effects can be estimated by using probabilistic estimation of residuals (PEER), an unsupervised method which builds off of factor analysis and has been shown to outperform PCA in identifying latent variables^15^.

After regressing out known and hidden covariates from the processed sequencing data in Section 3.1, we generated tables of QTLs: genomic variants whose genotype is significantly associated with gene expression (eQTLs), splicing (sQTLs), and chromatin accessibility (caQTLs) (Figure 1b) (see Methods, Figure S2a, Table S2-4).

Notably, regressing out these covariates began to reveal disease-relevant signals. Patients known to have C9-ALS cleanly separated from controls based on mRNA expression of the *C9orf72* gene, as expected (Figure 1c). We found 12 ALS lines that had not been labeled as C9-ALS in the clinical metadata but exhibited a pattern of low promoter accessibility resulting in reduced gene expression for *C9orf72* that mirrored that of the C9-ALS subjects; strikingly, the metadata labeled one of these lines as non-C9-ALS. Using a combination of ExpansionHunter^16^ and PCR, we found that these 12 samples, including the sample that had been labeled as non-C9-ALS in the clinical metadata, all indeed carried a mutant repeat expansion of at least 100 repeats. While there were no other non-C9 ALS lines that exhibited a similar pattern of C9orf72 expression downregulation, there was one control line (NEUYM205MRL) that exhibited C9orf72 expression levels comparable to those of C9-ALS patients, but not lower chromatin accessibility.

### 3.3 Characterization of cis-QTLs

QTL analysis demonstrated that most genes were influenced by at least one QTL within 1 Mbp (Table S2-4, Figure 2a-b), and that these QTL were consistent with the neuronal cell state and expected mechanisms of action. The slopes of significant eQTLs were highly correlated with the eQTLs from post-mortem brain tissue generated by the PsychENCODE consortium (Figure 2c)^17^. In comparisons to eQTLs generated from 48 tissues by the GTEx consortium^18^, our eQTLs were most similar to brain tissues in terms of correlation of effect sizes and number of gene-variant pairs in common (Figure S2b-c). Many genes were regulated by more than one QTL (Table S5-7, Figure S2e, see Methods). As expected, these higher order QTLs were located progressively farther away from omic features (example for eQTLs shown in Figure S2f).

**Figure 2.**
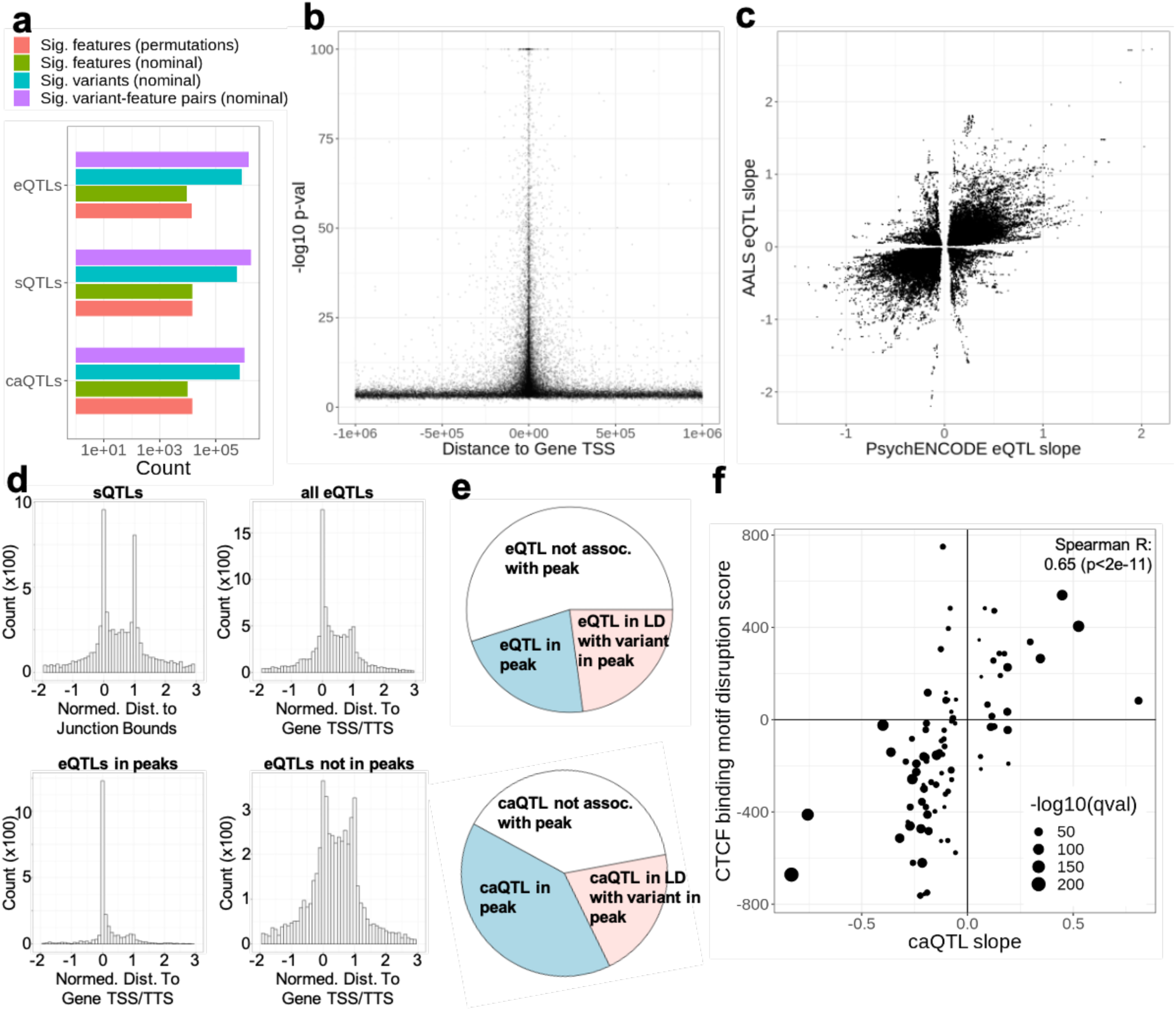
Characterization of identified QTLs. A) Number of significant QTLs at a Bonferroni corrected p<0.01. For eQTLs, features are quantifications of gene expression; for sQTLs, feature are quantification of intron splicing; for caQTLs, features are chromatin peak accessibilities. Colored lines represent numbers of significant variants and features in permutation and nominal analyses (see inserted legend, and methods for details). B) The -log10 p-value of all eQTLs as a function of the distance from the transcription start site of the associated gene. C) Comparison of slope from our analysis to PsychENCODE for all significant eQTL gene-variant pairs. D) Feature-length normalized distance to feature start sites for significant sQTLs and eQTLs. Top left: strand corrected, splice junction-length normalized distance to splice junction start. Top right: strand corrected, gene-length normalized distance to gene transcription start site (0 corresponds to TSS and 1 corresponds to TTS). Bottom right: same as top right, but subset to only eQTLs in peaks. Bottom left: same as top right, but subset to only eQTLs *not* in peaks. E) Pie charts denoting proportions of eQTLs and caQTLs in peaks, or in LD (linkage disequilibrium) with a variant in a peak. F) Motif disruption score plotted against caQTL slope for caQTLs falling in CTCF binding sites. Increase in circle size represents increase in -log 10 q value.

The positions of e/s/caQTLs relative to their target features (i.e. gene start, splice junction, chromatin peak) largely reflect the expected underlying regulatory mechanisms, with target features encompassing splicing variants typically near splice junctions, eQTLs near or in LD with variants near gene transcription start sites (TSS) and gene transcription termination sites (TTS), and caQTLs frequently near or in LD with variants in ATAC-seq peaks (Figure 2d-e). Variants were generally located within 10 kb of the omic feature they were most strongly associated with; most top eQTLs, sQTLs, and caQTLs were located within 5kb, 2.5kb, and 1 kb of their top feature, respectively (Figure S2d). Interestingly, caQTL effect sizes were highly correlated with motif disruption scores for CTCF binding (Figure 2f, see methods), but not other DNA binding proteins, emphasizing the central role of CTCF as a master chromatin regulator and the reproducibility of CTCF signals across diverse cell types.

### 3.4 Leveraging QTLs reveals consequences of ALS GWAS variants

We reasoned that our QTL analysis could help elucidate the molecular function of variants identified in prior GWAS. Indeed, more than half (970/1608) of the significant variants from Project MinE data (p<1e-5)^4^ were also a QTL; 753 loci were also an eQTL (with effects on 69 genes), 618 loci were also sQTLs (with effects on 81 genes), and 458 loci were also caQTLs (with effects on 30 genes) (Table S8-10). In total, variants had QTL effects on 106 genes. Among the most significant GWAS variants are those across the *C9orf72* locus, and these were also highly significant sQTLs and caQTLs for *C9orf72*.

Previously, Project MinE had reported an LD block of over 80 genomic variants spanning the *SCFD1* and *G2E3* genes as a novel risk locus for ALS, and all of these were also noted to be eQTLs for *SCFD1*^19^. Our results confirm that these variants are eQTLs for *SCFD1*; one of these variants, rs229143, falls within the SCFD1 promoter chromatin peak (Figure 3a-b). Importantly, we find that they are also sQTLs for *G2E3* exon 2 skipping; the only sQTL that falls within the introns adjacent to *G2E3* exon 2 is rs229190 (Figure 3b, Figure S3a). The omission of this exon is predicted to shift the open reading frame to exon 4. We note in passing that we also detected frequent exon skipping of *G2E3* exon 11 that was not significantly associated with genomic variation. A prior study examining truncated isoforms of the G2E3 protein^20^ suggests that exon 2 skipping would result in a truncated protein missing a nucleolar localization sequence, while exon 11 skipping would result in a truncated protein missing a nuclear localization sequence (Figure S3a).

**Figure 3.**
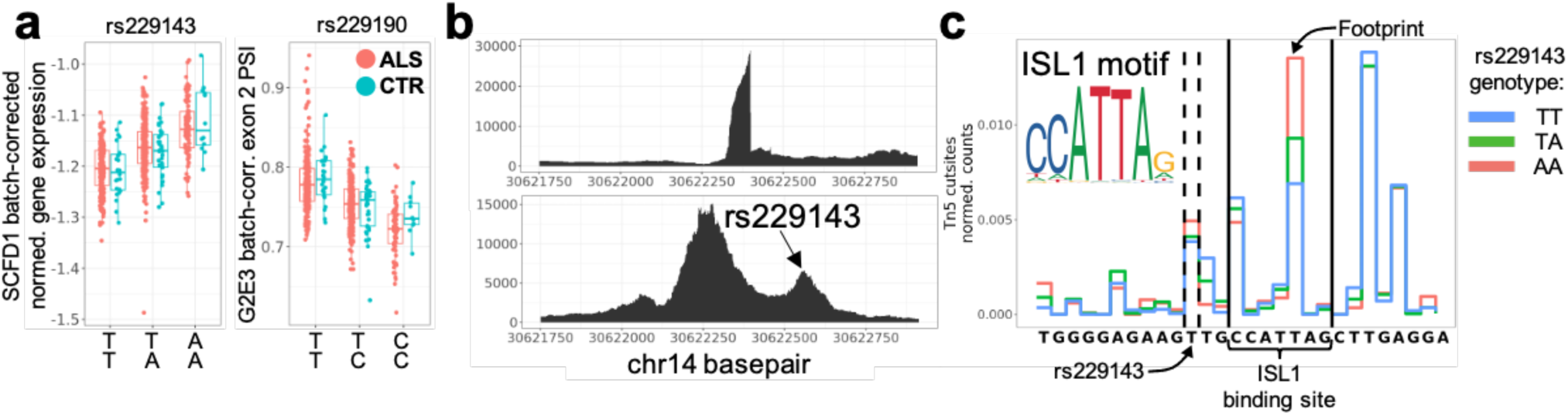
Overlapping QTL signals with Project MinE GWAS. A) Batch corrected expression of *SCFD1* plotted as a function of rs229143 genotype (adj. p-val<2e-32, b = 0.048+/-0.003) and batch corrected percent-spliced-in (PSI) of *G2E3* exon 2 as a function of rs229190 genotype (adj. p-val<6e-32, b = -0.029+/-0.002) for ALS (red) and CTR (blue). B) RNA-seq (top) and ATAC-seq (bottom) coverage of the same chromatin region corresponding to the SCFD1 promoter/exon 1 (promoter peak: chr14:30621205-30622913; exon 1: chr14:30622319-30622399). C) Peak-wise frequency of Tn5 cut sites across samples merged by rs229143 genotype. Variant between dashed lines corresponds to genomic location of rs229143. Region between solid lines corresponds to genomic location of JASPAR-predicted ISL1 binding site. Tn5 cut frequency at base pair labeled by arrow exhibits a strong dependency of rs229143 genotype. Color representation in insert.

Leveraging our ATAC-seq data and a transcription factor footprinting analysis^21^, we propose a causal mechanism for one of the 80 variants that may explain the role of this LD block. The variant rs229143, which is both a *SCFD1* eQTL (Figure 3a) and a significant GWAS variant, is located within an accessible chromatin peak in *SCFD1* intron 1 (Figure 3b). We hypothesized that rs229143 might disrupt a transcription factor binding site. To test this theory, we merged the aligned ATAC-seq reads at this locus by genotype and found a rs229143 genotype-dependent enrichment of Tn5 cuts in this region (Figure 3c). While the identity of the transcription factor cannot be determined by our data alone, we note that rs229143 is 2 bp upstream of the JASPAR-predicted^22^ binding site for the motor neuron specific marker ISL1 (Figure 3c). These results suggest a direct mechanistic link between rs229143 genotype, ISL1 transcription factor binding, and *SCFD1* gene expression; moreover, they highlight how multiple omics modalities can be integrated to derive mechanistic insights into disease.

### 3.5 *C9orf72* eQTLs are coinherited and form four C9-ALS patient subgroups with ordered disease progression rates

We next explored the possible role of QTLs in ALS caused by the mutant hexanucleotide repeat expansion (HRE) in the *C9orf72* gene (C9-ALS). Previous studies of gene expression in brains of people without ALS have found that common genomic variants at the *C9orf72* locus can alter *C9orf72* gene expression^17^. However, it is not clear if these eQTLs are relevant to C9-ALS.

We identified highly significant *C9orf72* sQTLs (corresponding to alternative 5’ splicing at exon 1a/1b) and eQTLs (Figure S3b), and less significant caQTLs (Table S11-13). Significant QTLs spanned the *C9orf72/MOB3B* locus and formed roughly a dozen linkage-disequilibrium (LD) blocks (correlation heatmap of a subset of significant QTLs is shown in Figure S3c). Although the local LD limits our ability to computationally identify causal variants, our analysis revealed that variation in *C9orf72* expression was most likely driven by the combined genotypes of two common variants (rs2492816, gnomAD MAF 0.41, and rs13691, gnomAD MAF 0.22) and one rare variant with a particularly large effect size (rs113860022, gnomAD MAF 0.011) (Figure 4a, Figure S3d). The variant rs13691 falls within the 3’UTR of *C9orf72* and likely disrupts mRNA stability, while rs113860022 falls within the promoter peak of *C9orf72* and likely disrupts transcription factor (TF) binding (Figure 4b). The previously mentioned control line that exhibited *C9orf72* expression levels comparable to C9-ALS cases (NEUYM205MRL, Section 3.2) was a homozygous carrier for the alternate allele of rs113860022. This donor was 60 years old at sampling, only slightly older than the reported median age at onset for C9orf72-associated ALS (∼58 years)^23^, so it is possible that this individual carried a pathogenic variant but had not yet developed symptoms.

**Figure 4.**
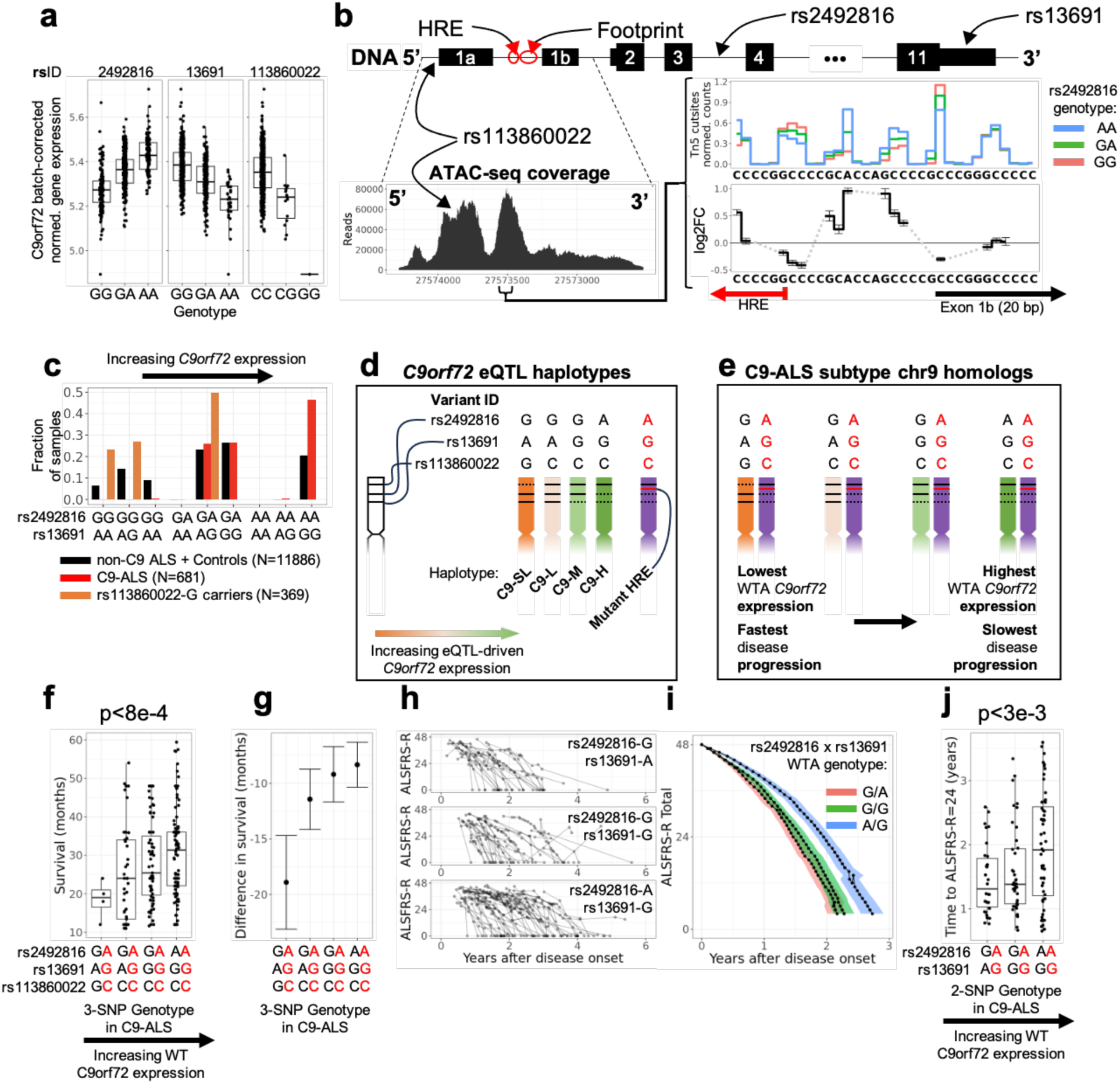
C9orf72 QTLs are coinherited and form four C9-ALS subtypes. A) *C9orf72* batch-corrected normalized gene expression plotted as a function of rs2492816 (adj. p-val<2e-37, slope = 0.086 +/- 0.006), rs13691 (adj. p-val<9e-23, slope = -0.081 +/- 0.007), and rs113860022 (adj. p-val < 3e-3, slope = -0.16 +/-0.02). Genotypes plotted in order of homozygous reference allele, heterozygous, homozygous alternate allele, as defined by dbSNP. B) Schematic showing the location of top *C9orf72* QTLs and the *C9orf72* promoter peak, as measured by ATAC-seq. Inset showing normalized Tn5 cutsite counts at single basepair resolution, as a function of genotype. Log2FC corresponds to change in cutsite counts per rs2492816-A allele (0 for rs2492816-GG, 1 for rs2492816-GA, 2 for rs2492816-AA, see Table S16). Error bars correspond to DESeq2 standard errors. Gray dashes serve as visual cues to connect basepairs with sufficiently high counts for differential analysis; gray dashes span basepairs excluded from differential analysis due to low Tn5 cut counts (less than 1 detected Tn5 cut per sample on average). C) Population frequencies of rs2492816 /rs13691 2-SNP genotypes in the ALS Compute database. Values on the y axis denote fractions of samples in each group; black: non-C9 ALS and controls; red: C9-ALS patients; orange: non-C9 ALS and controls heterozygous for rs113860022. D) *C9orf72* eQTL haplotypes that can be inferred from population frequency data in (C); C9-SL: C9-super-low; C9-L: C9-low; C9-M: C9-medium; C9-H: C9-High. The mutant-HRE is coinherited with C9-H, but exhibits the lowest expression. E) Cartoon illustrating the four eQTL subgroups of C9-ALS patients that we identify. All eQTL genomic variation is restricted to the wild-type-allele (WTA). Labels and colors match (D). F) Disease length in C9-ALS patients from the ALS Compute cohort plotted as a function of the combined genotypes of rs2492816 (top genotype), rs13691 (middle genotype), and rs113860022 (bottom genotype). Red letters indicate mutant-HRE allele genotype, and black letters indicate implied genotype of wild-type allele based off of haplotype analysis (see D-E). P-value calculated on log transformed data to improve normality of residuals. Top and bottom 5% of data points were removed (agnostic of group) prior to regression to reduce the effect of outliers. G) Mean difference in survival between C9-ALS patients and non-C9-ALS patients as a function of C9-ALS subgroup genotype. For each 3-SNP genotype, we used a t-test to compare the difference in survival between C9-ALS subjects and non-C9-ALS subjects with the same genotype; the depicted summary statistics correspond to the outputs of this t-test. Error bars denote standard error. Raw data depicted in Figure S3e. Genotypes are ordered as in panel (E). Red letters indicate mutant-HRE allele genotype, black letters indicate wild-type allele genotype. H) Longitudinal ALSFRS-R total as a function of time from disease onset for C9-ALS patients carrying the wild-type allele 2-SNP genotype of rs2492816-G/ rs13691-A (top), rs2492816-G/ rs13691-G (middle), and rs2492816-A/ rs13691-G (bottom). No ALSFRS-R progression data was available for carriers of the rs113860022-G allele. I) Genotype-averaged ALSFRS-R trajectories from (H); ribbons denote standard error. Point-wise estimates of time-to-ALSFRS-R Total statistics can be found in Figure 4j and Figure S3g. J) Time to ALSFRS-R =24 from time of disease onset, plotted as a function of the combined genotypes of rs2492816 (top genotype) and rs13691 (bottom genotype) for C9-ALS patients. Red letters indicate mutant-HRE allele genotype, and black letters indicate implied genotype of wild-type allele based off of haplotype analysis (see Figure 4e). Top and bottom 5% of data points were removed (agnostic of group) prior to regression to reduce the effect of outliers.

The functional role of rs2492816, which is in intron 3, was less clear (Figure 4b, top). Observing that rs2492816 is both the most highly significant sQTL and eQTL for *C9orf72*, we hypothesized that it may disrupt the binding of pioneer transcription factors, thereby affecting recruitment of other TFs at the *C9orf72* promoter. We indeed found that the rs2492816-A allele is significantly associated with a higher local ATAC-seq signal immediately downstream of the HRE and roughly 20 bp upstream of the V2 transcript promoter (chromosome-wide significance p<1e-40), lending support to our hypothesis (Figure 4b, inset), likely indicating a disrupted protein-DNA interaction (see Discussion).

To better understand the relevance of these genomic variants to C9-ALS, we determined whether the SNPs were associated with the wild-type or mutant-HRE allele. Using data from AALS (n=956) and the ALS Compute database (n>12,000), we found that variation at rs2492816, rs13691, and rs113860022 in C9-ALS patients is restricted to the wild-type allele (WTA). The unphased genotype frequencies of the two common variants (rs2492816 and rs13691, Figure 4c) could be explained by three founder haplotypes, each with a distinct *C9orf72* expression profile: rs2492816-G/rs13691-A (C9-low), rs2492816-G/rs13691-G (C9-medium), and rs2492816-A/rs13691-G (C9-high) (Figure 4d). Carriers of rs2492816-A/ rs13691-A were notably missing, suggesting a historical bottleneck. Interestingly, the rare allele rs113860022-G was consistently co-inherited with, and formed a subset of, the C9-low haplotype (rs2492816-G/ rs13691-A), forming a C9-“super-low” group (Figure 4d).

Importantly, we also found that the *C9orf72* mutant HRE (AALS n=50, ALS Compute n>600) was co-inherited with, and formed a subset of, the C9-high haplotype (rs2492816-A/ rs13691-G) (Figure 4c-d). This is in line with previous findings that most mutant HREs are observed on the “Finnish founder haplotype”^7,24,25^. Together, these results enable the stratification of all C9-ALS patients into four genotype-defined groups with clearly ordered *C9orf72* expression levels, determined by their WTA haplotype (Figure 4e).

We sought to determine whether these four genotype-defined groups, which drive *C9orf72* expression changes through variation on the WTA, can alter disease progression in C9-ALS. Within the C9-ALS cohort of ALS Compute, we found a significant association (p<8e-4) between disease duration and the WTA haplotype for 218 C9-ALS patients (Figure 4f); the wild-type C9-high haplotype exhibited a protective effect, while the C9-super-low group notably exhibited the fastest disease progression. At the same time, no genotype-dependent trend was detected in non-C9-ALS patients (Figure S3e).

To confirm that the WTA haplotype was modifying the effects of the mutant allele, we compared the difference in survival between C9- and non-C9 ALS patients in ALS Compute who shared the same WTA haplotype (Figure 4g, S3e). Notably, C9-ALS patients carrying the C9-high haplotype on their WTA had a 8+/-2 month (standard error, p < 6e-5) shorter survival than non-C9 ALS patients with the same genotype, whereas C9-ALS patients carrying the C9-super-low haplotype on their WTA had a 18 +/- 5 month (standard error, p < 3e-4) shorter survival than non-C9 ALS patients. (Figure 4g, S3e).

Given the uncertainty of disease duration data, we conducted a second analysis using longitudinal data of ALSFRS-R scores gathered from AALS and from the Emory clinic (Figure 4h-j). For each of the 112 C9-ALS patients, we estimated the time from disease onset for a patient to reach an ALSFRS-R score of 24 using linear interpolation (Figure S3f, see methods). Even though the dataset was smaller (and there were no carriers of the C9-super-low haplotype), we found a similarly significant association between the time to ALSFRS-R=24 and the *C9orf72* WTA haplotype for C9-ALS patients (p<3e-3) (Figure 4j). A range of other ALSFRS-R thresholds were tested to verify the robustness of our method (Figure S3g). In summary, disease progression in C9-ALS patients is associated with SNPs that modify expression of the *<u>wild-type</u>*allele of *C9orf72*.

### 3.6 Identifying functional effects of rare variants reveals novel cryptic exons in ALS genes

Several known ALS genes carry rare loss of function (LOF) variants that were not detectable by GWAS^4,26–29^. We hypothesized that additional pathogenic variants, ranging from SNPs to genomic deletions, might exist that act by dramatically reducing expression of known ALS genes. Indeed, we identified individuals exhibiting severely downregulated expression (>3 interquartile ranges below the cohort median) in *NEK1* (6 samples), *OPTN* (3 samples), *TBK1* (2 samples), *SETX* (2 samples), *SPG11* (3 samples), and *ATP13A2* (1 sample) (Figure 5a) (each mutation was detected in only one sample). Of these 17 cases, 3 carry large genomic deletions, 8 carry exonic stop-gain mutations, and 6 carry splice-altering mutations (Figure 5b, caption). In these cases, the mutations likely induce nonsense-mediated decay resulting in anomalously low expression.

**Figure 5:**
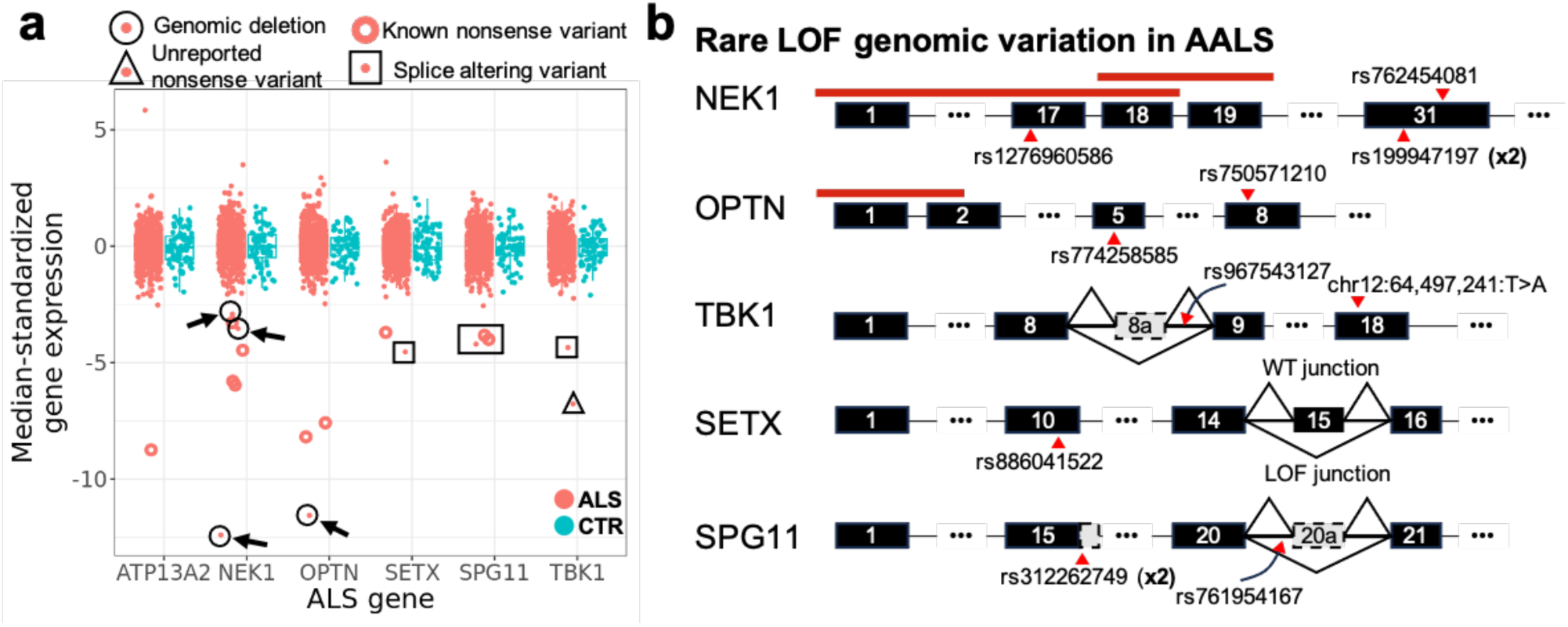
Matching rare LOF mutations to expression phenotypes. A) Median-zeroed, IQR-standardized expression of six ALS genes. Red unfilled circles correspond to samples that carry a heterozygous mutation for either a known nonsense mutation or a frameshift variant that leads to an early stop. Points surrounded by a black circle and an arrow carry a heterozygous deletion of a coding portion of the gene. Point surrounded by a triangle carries a nonsense mutation that is not in dbSNP. Points surrounded by boxes correspond to samples which exhibit a deleterious splicing pattern. B) Schematic of gene mutations found in A. Red lines indicate spans of deletions (black circles with arrows in A). Multipliers (x2) indicate mutations that occurred in two samples. *ATP13A2* is excluded because the single sample with downregulated expression can be explained by an exonic frameshift mutation (rs765632065).

Surprisingly, all but two causal SNPs appeared in only one sample each, highlighting the fact that there are many different genetic causes that can result in the same transcriptomic phenotype. Of the variants in Figure 5b, six have not been previously reported as associated with ALS in CLINVAR^30^ or a recent study annotating pathogenic variants in sporadic ALS^31^, but carry the same functional consequence as variants that have. Previously unreported variants include a nonsense mutation in *TBK1* that has not been reported in dbSNP,^32^ an ultrarare intronic variant (rs967543127) in *TBK1* which we link to a retained cryptic exon downstream of exon 8 (Figure S4a), and an ultrarare intronic variant in *SPG11* (rs761954167) which we link to a retained cryptic exon downstream of exon 20 (Figure S4b).

In contrast to the previous cases, we were unable to identify a mutation that explains the downregulation of *SETX* for NEUZX847VWV. Nevertheless, it is likely that the downregulation of this gene is allele-specific, as NEUZX847VWV exhibits the most imbalanced RNA-seq allele coverage of all 594 samples across four heterozygous mutations in exon 10 (Figure S4c). In this individual, the out-of-frame exon 15 is skipped much more frequently than in other samples, which induces an early stop codon (Figure 5b, Figure S4d). While the sample was not a carrier of any unique biallelic SNPs or small indels between exon 14 and 16, this does not preclude the existence of a more complex causal genomic variant at that locus.

## 4 Discussion

In this study, we assembled a QTL atlas from iMNs generated from 594 ALS patients and healthy controls. By integrating ATAC-seq, RNA-seq, and WGS, we identify regulatory relationships between omic modalities, and signals that are most likely relevant to ALS. We identify regulatory variation at the *C9orf72* locus that modifies disease progression in C9-ALS, and uncover rare, often noncoding or structural variants that likely explain disease in additional patients who previously lacked a molecular diagnosis.

### 4.1 Common variants can alter disease progression in C9-ALS

We found that common genomic variants are significantly associated with altered disease progression in C9-ALS. The variants we identified had effect sizes of 6 to 12 months, which is on par or greater than the survival extension by the ALS drugs riluzole and edaravone^1^. As such, this finding has implications for patient stratification for clinical trials. The mechanism of action of these variants is perhaps somewhat unexpected. Previous studies have primarily searched for disease-associated changes related to the mutant allele of *C9orf72*^33–37^ . By contrast, the disease-modifying variants that we detect are located on the healthy, wild-type chromosome that alter expression of the wild-type *C9orf72* allele, with higher levels of the wild-type *C9orf72* associated with significantly slower disease progression. This finding raises an intriguing possibility that disease-modifying eQTL or sQTL variants may be found for other causes of ALS by looking for loci that boost expression of the wild-type allele to partially compensate for the mutant allele. Moreover, the observations raise intriguing possibilities for future therapeutic strategies based on such compensatory mechanisms.

There are precedents in the literature for the possibility that higher levels of wild-type *C9orf72* compensate for the mutant allele. While the mutant HRE can induce *C9orf72* haploinsufficiency^7,38–40^, C9-ALS pathology is thought to be primarily driven by accumulation of toxic RNA foci and dipeptide repeats derived from the mutant *C9orf72* HRE. The compensatory mechanism we observe may be related to the role of C9orf72 in cellular autophagy, the process responsible for eliminating these toxic species^41,42^. Previous work has shown that exogenous *C9orf72* downregulation can exacerbate the accumulation of these toxic species. In particular, Boivin et al. have shown that partial siRNA-mediated knockdown of *C9orf72* results in the increased accumulation of polyPG, polyGA, and polyGR dipeptide proteins in GT1-7 neuronal cells, which can be partially rescued with drugs activating autophagy^42^. Zhu et al. have shown that transgenic mice expressing the mutant HRE exhibited early death and cognitive deficits in response to *C9orf72* gene deactivation^41^. At the same time, Liu et al. demonstrated that the survival of C9HRE iMNs under tunicamycin-induced stress could be increased by exogenous expression of C9orf72 protein^43^. In the context of our findings, it is likely that the endogenous, naturally-occurring genome-driven upregulation of wild-type allele *C9orf72* in eQTL carriers results in higher protein expression, which in turn improves autophagy and clearance of toxic DPRs, thereby delaying disease progression, providing genetic evidence in support of therapeutic strategies aimed at increasing autophagy in C9-ALS.

Our data are also consistent with a pure loss of function mechanism of neurodegeneration due to reduced autophagy/lysosomal function^7^ in both neurons and microglia^44^. Notably, Biogen conducted a clinical trial of an antisense oligonucleotide (ASO) targeting C9ORF72 which succeeded in lowering the levels of the gene and significantly reducing the DPRs that were measured; nevertheless, it had no beneficial clinical effect^45^. C9-ALS/FTD is also associated with TDP-43 pathology in neurons^46^, and it is possible that haploinsufficiency of C9orf72 alone is sufficient to cause impairment of autophagy/lysosomal function and neuroinflammation. Relatedly, mutations leading to haploinsufficiency of progranulin, a gene also involved in autophagy and lysosomal function, have been found to cause FTD, but without generating DPRs or RNA foci^47,48^. Thus, in neurons, C9orf72 haploinsufficiency could cause TDP-43 pathology and neurodegeneration; in microglia, it could cause a loss of its trophic role for neurons and a propensity to become chronically inflamed, which would exacerbate neurodegeneration.

Relatedly, the study of Liu et al.^43^ also offers a candidate protein for the putative rs2492816-dependent DNA-protein interaction in the *C9orf72* V2 promoter (Figure 4b). The ATAC and RNA signals associated with rs2492816-A (Figure 4a-b) suggest that it is associated with the disrupted binding of a transcriptional repressor. One candidate is the epigenetic repressor DAXX, which Liu et al. found to bind the G4C2 repeat and selectively alter transcription at the *C9orf72* V2 promoter^43^. This raises the possibility that the rs2492816 eQTL may additionally play a role in the DNA toxicity hypothesis for C9-ALS pathogenesis proposed by their study.

### 4.2 Integration of ATAC-seq, RNA-seq, and WGS reveals ALS-associated signals

In parallel with these modifier effects, we identified several rare genomic variants which contribute to loss of function in known ALS genes; as these variants mirror the effect of mutations known to contribute to ALS pathology, it is possible that they contribute to patient-specific causes of disease as well. This is especially the case for TBK1, a kinase that plays a major role in cell autophagy, loss of function mutations in which have been found to cause ALS with autosomal dominant inheritance^49^. Heterozygous ALS-related loss of function mutations have also previously been described in NEK1, a kinase implicated in cell cycle regulation, axonal development, and DNA damage repair, and OPTN, a cargo protein that is phosphorylated by TBK1 and also plays an important role in cell autophagy; however, the mutation penetrance is unclear^50–53^. Mutations across SPG11, a gene involved in the lysosomal formation^54^, and ATP13A2, a lysosomal ATP-ase^55^, have both been found to cause ALS with autosomal recessive inheritance^56,57^. Homozygous loss of function mutations in both of these genes have been found to cause juvenile onset ALS^56,57^, suggesting that gene downregulation could be a risk factor for disease; given the observation that all the deleterious SPG11 variants we detect are intronic splice altering variants, it is possible that mutations in this gene are under-reported due to the difficulty in detecting and accurately assessing their pathogenic burden. Mutations in ATP13A2 have also been found to cause an autosomal recessive inherited version of Parkinson’s disease; heterozygous loss of function mutations have been found within the gene as well, and it has been suggested that they serve as a risk factor for sporadic PD^58^. We also identified heterozygous loss of function phenotypes within the SETX gene, a helicase that plays a role in autophagy, the DNA damage response, and R-loop resolution^59^. Mutations in SETX cause ALS by a gain-of-function mechanism with juvenile disease onset, while homozygous loss of function mutations in SETX have been found to cause ataxia with oculomotor apraxia type 2^60,61^. This raises the possibility that nonsense mutations in SETX can contribute to sporadic ALS risk. Interestingly, the individual that exhibited disrupted splicing of SETX also carried a heterozygous nonsense mutation in the ATM gene which plays a key role in the ATM-DNA damage response which repairs double-strand breaks^62^; relatedly, a previous study showed that hypoxia-induced replication stress in RKO cell lines induces both the ATM-dependent DNA damage response and SETX expression as protective effects^63^ (PMID: 34140498). While neither of these mutations are definitive causes for disease by themselves, it is conceivable that they both contribute to the dysfunction in a larger common pathway, culminating in ALS pathology.

Notably, over half of the novel, rare genomic variants that we identify were unlikely to have been detected by standard rare-variant analysis. Three samples carried kilobase-scale deletions in the *NEK1* and *OPTN* genes, and 6 samples carrying intronic splice-altering variants in the *TBK1*, *SETX*, and *SPG11* genes. Large deletions are often excluded from traditional burden tests due to their heterogeneity in location and size, and intronic variants are typically excluded as long as they fall over 2bp away from an exon, precluding their label as a potential splice variant. If representative, these results suggest that a large share of ostensibly unexplained ALS cases carry hard-to-detect variants in established ALS genes, bridging the gap between the relatively small fraction (15%) of ALS patients with known inherited mutations and the much higher estimated heritability of disease (50%).

## 5 Conclusion

Overall, our results show the impact of common genetic regulatory variants on disease progression in C9-ALS and identify rare genetic events that can directly account for disease in individual patients. Among our cohort of 522 patients, 58 had clinical data indicating that they carry previously recognized disease-causing mutations in known ALS genes. Our multi-omic analysis revealed that an additional 17 individuals had disease that was likely caused by eQTL or sQTL variants targeting known ALS genes. Extrapolating from this study, it seems plausible that the fraction of ALS patients whose disease is attributable to known ALS genes may be substantially higher than previously recognized. Conversely, our findings also suggest that there may be many extremely rare causes of ALS that will be highly challenging to find using conventional methods.

## Methods

### Patient demographics

Overall, clinical data showed that 63 individuals with ALS had a family history of the disease. This is likely an underestimate, though, as no family history was recorded for 88 individuals, and either a paternal or maternal record was entirely missing for 137 individuals. Only 234 individuals had at least one maternal and one paternal record and did not have a family history of ALS (Figure S1e). Out of the 36 individuals that carried the mutant *C9orf72* HRE, only half (18) had family history of disease (Figure S1f). All of the 12 individuals that carried the SOD1 mutation had a family history of disease. Only one control had a family history of ALS.

### Generation of iPSC motor neuron cultures, whole genome sequencing data, RNA-seq data, and ATAC-seq data

Methods for generating iPSC motor neuron cultures, whole genome sequencing data, and RNA-seq data were published previously^6,8,9^. Data was downloaded from the Answer ALS dataportal.

### Splicing processing

Alternative splicing was quantified using LeafCutter (v0.2.9), following the recommended workflow. First, raw reads were aligned to the GRCh38 using STAR (--twopassMode Basic, --outSAMstrandField intronMotif). Then splice junctions were extracted from the STAR alignments using regtools (junctions extract) with parameters chosen to retain high-confidence junctions and reduce short overhang artifacts: -a 8 -m 50 -M 500000 -s RF. Junction files from all samples were then used as input for intron clustering with LeafCutter. Intron clusters were generated using the LeafCutter regtools-compatible clustering script (leafcutter_cluster_regtools.py) with the following parameters: -m 50 -l 500000, producing cluster definitions were used for downstream splicing analyses.

### Generation of PEER factors

PEER factors were generated using the “peer” package in R using default settings^15^. For each omics matrix (vst-normalized gene counts, vst-normalized peak counts, and leafcutter PSI matrix) we generated 100 PEER factors. PEER factors were found to correlate with known covariates (S100B staining, Nestin staining, FRiP score) and across omics modalities. Top genes affected by regressing out PEER factors were enriched for transcription factors (nominal p-value 1e-12).

### QTL summary statistics

QTLs were generated using the tensorQTL Python package.^64^ Baseline covariates for each omic are listed as follows. Gene expression: sex, iPSC cell type of origin, and ALS case status. Splicing: sex, iPSC cell type of origin, and ALS case status. Chromatin accessibility: sex, iPSC cell type of origin, FRiP score, sequencer, and ALS case status. All covariates except for FRiP score were binary. We additionally included PEER factors as covariates for each omic modality to account for hidden batch effects. We chose the number of PEER factors as covariates that maximized the number of phenotypes significantly associated with at least one genomic variant. Specifically, for each omics modality, we reran tensorQTL in the “cis” mode 21 times, each time adding an additional 5 PEER factors as covariates at each run (run 1: 0 PEER factors, run 2: 5 PEER factors, … run 21: 100 PEER factors) and recording the total number of significant phenotypes each time (Figure S2a). We generated our final summary statistics for this study using 50 PEER factors for gene expression, 10 for splicing, and 20 for chromatin accessibility. For each omic modality, we generated summary statistics using the “cis” and “cis_nominal” mode of tensorQTL for all variant-phenotype pairs within a 1 Mb window (of splice junction center for splicing, peak center for chromatin accessibility, and transcription start site for gene expression). We also generated a table of conditionally independent QTLs using the tensorQTL “cis_independent” mode.

### QTL comparisons with GTEx and PsychENCODE data

Summary statistics for PsychENCODE^17^ and GTEx^18^ were downloaded from their respective websites. After intersecting variant-phenotype pairs, we calculated Pearson correlation coefficients between eQTL slopes of our dataset and the GTEx and PsychENCODE datasets. We only retained variant-phenotype pairs from our dataset that exhibited a Bonferroni-corrected p-value of 1e-2.

### Estimating motif disruption scores

The motif disruption score database was downloaded from SNP2TFBS^65^. Significant QTLs were intersected with variants predicted to disrupt transcription factor binding. For each transcription factor, the motif disruption score from SNP2TFBS was correlated with QTL slope (for intersecting variants). Only CTCF was found to be significant.

### Generating single-basepair ATAC-seq signal

To make coverage plots of ATAC-seq at single basepair resolution, we tabulated the number of Tn5 cuts per base per sample after shifting reads using the standard +5/-4 bp shift using Rsamtools^66^ to generate a base by sample counts matrix for each peak of interest. For genotype specific plotting (Figure 3c, 4b inset), we aggregated by summing across samples from the same genotypes, and we normalized samples by total cuts in that peak.

### Identifying *C9orf72* QTLs

We conducted a fine mapping analysis to identify conditionally independent variants that alter C9orf72 expression using the SusieR^67^ package. As ALS patients carrying the mutant HRE are known to exhibit downregulated *C9orf72* expression, we only included samples that did not carry the mutant C9orf72 HRE in this analysis. We searched for variants within 10 kb of the C9orf72 gene body. We fit the data using a coverage of 0.99, and identified three credible sets. The first two credible sets consisted of one variant each: rs2492816 and rs113860022. The variant rs2492816 was also the top eQTL found in a study of post-mortem brain tissue^16^. The third credible set consisted of six variants, indicating that all of these variants are equally likely to be the causal variant; as rs13691 was the only variant of these six to fall within an exon of *C9orf72*, we selected it as the most likely causal variant for its likely functional role in the 3’ UTR of *C9orf72*.

### Correlating rs2492816 with *C9orf72* exon 1b promoter footprint

For each peak across chromosome 9, we tabulated the number of Tn5 cuts per base per sample, to generate a base by sample counts matrix for each peak. All matrices were stacked, and then rows with less than an average of 1 count per sample were removed. The remaining matrix was treated as a raw counts matrix and fed into DESeq2^68^ using sex, FRiP score, and rs2492816 genotype as covariates. Top significant hits (adj. p-val<1e-40) were found to fall in the promoter of exon 1b of *C9orf72*. Plotted Log2FC and error bars reflect DESeq2 log2FC and standard errors (Figure 4b).

### Correlating C9-ALS subgroups with disease length

In the ALS Compute database, we retrieved information on all C9-ALS patients (sample IDs in Table S14) with known disease survival and subset to either limb or bulbar onset. Prior to subgroup-specific analysis, patients with the top 5% longest survival and bottom 5% shortest survival were removed. Due to irregularity of residuals, a linear model (*lm* function in base R) was used to fit log10 (survival) to numerically encoded C9-ALS subgroup (rs2492816/rs13691/rs113860022 genotypes were encoded as follows: GA/AG/GC - 0, GA/AG/CC - 1, GA/GG/CC - 2, AA/GG/CC - 3). The p-value in Figure 4f reflects the fit of this model.

### ALSFRS-R Total data correlations

ALSFRS-R Total data for C9-ALS patients with genotype data at rs2492816 and rs13691 was collected from Answer ALS and an ALS Clinician (sample IDs in Table S15). We initially attempted to conduct an analysis with ALSFRS-R slope, but we found that this metric had no correlation with length of disease, confirming previous assertions that ALSFRS-R Total slope is a noisy metric for studying disease progression rates. Instead, we decided to use an alternate approach: estimating the time from disease onset to the time for the patient to reach an ALSFRS-R Total value of 24 (half of the maximum, 48). As such, only samples with a known date of disease onset were retained. ALSFRS-R total score at date of disease onset was anchored at 48. ALSFRS-R total score at date of death (if known) was anchored at 0. ALSFRS-R total score between known values was estimated using linear interpolation as shown in Figure S3f. Average progression plots in Figure 4i were constructed as follows: for each ALSFRS-R Total score *i,* and each patient *j*, the time from disease onset to reach score *i*, t*i,j*, was estimated using linear interpolation; patients with the top 5% longest times and bottom 5% shortest times were removed agnostic of genotype; mean values and standard errors of the t*i,j* values were then computed across patients within each genotype group to generate the average progression plots shown in Figure 4i. The data in Figure 4h reflects the interpolated times to reach an ALSFRS-R score of *i=24*. To ensure that the significance we detected was not an artifact, we computed the p-value for a range of thresholds spanning ALSFRS-R Total of 3 to 45, as shown in Figure S3g.

### Outliers in ALS genes

To identify outliers in known ALS genes, we searched for samples with anomalously low expression (batch-corrected expression >3 interquartile ranges below the median) in known ALS genes. For each anomalous sample, we manually inspected genomics and transcriptomics read coverage to identify causes. Structural variants were found from manual inspection of genomics data and confirmed using Delly^69^. All unique splicing events were analyzed in the context of all samples to ensure uniqueness.

## Data Availability

All data not included in the supplementary information of this study, including RNA-seq counts matrices, ATAC-seq counts matrices, and splicing percent-spliced-in fractions can be downloaded from the Answer ALS dataportal. Full feature-variant QTL summary statistics have been made available on the Answer ALS dataportal as well. Longitudinal ALSFRS-R Total data from Answer ALS is available with the clinical data and downloadable from the Answer ALS dataportal. Longitudinal ALSFRS-R Total data from the Emory clinic is available from the authors upon reasonable request. Clinical and genomics data from ALS Compute can be downloaded following instructions on dbGaP under accession phs003184.

## Consortia

Additional members of the Answer ALS Consortium are Nhan Huynh^1^, Danielle Firer^1^, Velina Kozareva^1^, Aneesh Donde^1^, Connie New^1^, Michael J. Workman^8^, Dhruv Sareen^26^, Loren Ornelas^26^, Lindsay Panther^26^, Erick Galvez^26^, Daniel Perez^26^, Imara Meepe^26^, Viviana Valencia^26^, Emilda Gomez^26^, Chunyan Liu^26^, Ruby Moran^26^, Louis Pinedo^26^, Richie Ho^8^, Terri Thompson^14^, Dillon Shear^14^, Robert Baloh^8^, Maria G. Banuelos^8^, Veronica Garcia^8^, Ronald Holewenski^7^, Oleg Karpov^7^, Danica-Mae Manalo^7^, Berhan Mandefro^8^, Andrea Matlock^7^, Rakhi Pandey^7^, Niveda Sundararaman^7^, Hannah Trost^8^, Vineet Vaibhav^7^, Vidya Venkatraman^7^, Oliver Wang^7^, Arish Jamil^15^, Naufa Amirani^4,5^, Leandro Lima^4,5^, Krishna Raja^4,5^, Wesley Robinson^4,5^, Reuben Thomas^4,5^, Edward Vertudes^4,5^, Stacia Wyman^4,5^, Carla Agurto^16^, Guillermo Cecchi^16^, Raquel Norel^16^, Omar Ahmad^9,10^, Emily G. Baxi^9,10^, Aianna Cerezo^26^, Alyssa N. Coyne^9,10^, Lindsey Hayes^26^, John W. Krakauer^26^, Nicholas Maragakis^26^, Elizabeth Mosmiller^26^, Promit Roy^26^, Steven Zeiler^26^, Miriam Adam^1^, Tobias Ehrenberger^1^, Alex Lenail^1^, Natasha Leanna Patel-Murray^1^, Yogindra Raghav^1^, Divya Ramamoorthy^1^, Karen Sachs^1^, Brook T. Wassie^1^, James Berry^17^, Merit E. Cudkowicz^17^, Alanna Farrar^17^, Sara Thrower^17^, Sarah Luppino^17^, Lindsay Pothier^17^, Alexander V. Sherman^17^, Ervin Sinani^17^, Prasha Vigneswaran^17^, Hong Yu^17^, Jay C. Beavers^18^, Mary Bellard^19^, Elizabeth Bruce^19^, Senda Ajroud-Driss^20^, Deniz Alibazoglu^20^, Ben Joslin^20^, Matthew B. Harms^21^, Sarah Heintzman^21^, Stephen Kolb^21^, Carolyn Prina^21^, Daragh Heitzman^22^, Todd Morgan^22^, Ricardo Miramontes^11^, Jennifer Stocksdale^23^, Keona Wang^23^, Jennifer Jockel-Balsarotti^24^, Elizabeth Karanja^24^, Jesse Markway^24^, Molly McCallum^24^, Tim Miller^24^, and Jennifer Roggenbuck^25^.

^14^On Point Scientific Inc., San Diego, CA, USA. ^16^Computational Biology Center, IBM T.J. Watson Research Center, Yorktown Heights, NY, USA. ^17^Department of Neurology, Healey Center, Massachusetts General Hospital, Harvard Medical School, Boston, MA, USA. ^18^Microsoft Research, Microsoft Corporation, Redmond, WA, USA. ^19^Microsoft University Relations, Microsoft Corporation, Redmond, WA, USA. ^20^Department of Neurology, Northwestern University, Chicago, IL, USA. ^21^Department of Neurology and Genetics, Ohio State University Wexner Medical Center, Columbus, OH, USA. ^22^Texas Neurology, Dallas, TX, USA. ^23^Department of Psychiatry and Human Behavior and Sue and Bill Gross Stem Cell Center, University of California, Irvine, CA, USA. ^24^Department of Neurology, Washington University, St. Louis, MO, USA. ^25^Zofia Consulting, Reston, VA, USA. ^26^Cedars-Sinai Biomanufacturing Center, Cedars-Sinai Medical Center, Los Angeles, CA, USA.

## Acknowledgements

Support for the Answer ALS program was kindly provided by the Robert Packard Center for ALS Research at Johns Hopkins, Travelers Insurance, ALS Finding a Cure Foundation, Stay Strong Vs. ALS, Answer ALS Foundation, Microsoft, Caterpillar Foundation, American Airlines, Team Gleason, National Institutes of Health, Fishman Family Foundation, Aviators Against ALS, AbbVie Foundation, Chan Zuckerberg Initiative, ALS Association, National Football League, F. Prime, M. Armstrong, Bruce Edwards Foundation, The Judith and Jean Pape Adams Charitable Foundation, Muscular Dystrophy Association, Les Turner ALS Foundation, PGA Tour, Gates Ventures, and Bari Lipp Foundation. This work was also supported in part by grants from the California Institute for Regenerative Medicine. Additional support for this work was provided by EverythingALS and the Vision 2030 Hub initiative.

This work was supported in part by the Intramural Research Programs of the NIH, the National Institute on Aging (ZIA-AG000935), and the National Institute of Neurological Disorders and Stroke (ZIA-NS003154). Additional collaborators, sources of support, and origin of the data and biospecimens are listed in the following publications: (1) Chia R, Sabir MS, Bandres-Ciga S, Saez-Atienzar S, Reynolds RH, Gustavsson E, et al. Genome sequencing analysis identifies new loci associated with Lewy body dementia and provides insights into its genetic architecture. Nat Genet. 2021 Mar; 53(3):294-303. PMID: 33589841; and (2) Dewan R, Chia R, Ding J, Hickman RA, Stein TD, Abramzon Y, et al. Pathogenic Huntingtin Repeat Expansions in Patients with Frontotemporal Dementia and Amyotrophic Lateral Sclerosis. Neuron. 2021 Feb 03; 109(3):448-460.e4. PMID: 33242422.

We would like to acknowledge sample and data contributions from the “Genomic Translation for ALS Care” project investigators and funding sources: PI Harms, Matthew (Columbia University) and site investigators Appel, Stanley (Houston Methodist); Baloh, Robert (Cedars Sinai); Bedlack, Richard (Duke Univ); Chandran, Siddharthan (Univ. of Edinburgh); Foster, Laura (Univ of Colorado); Gibson, Summer (Univ Utah); Goldstein, David (Columbia University); Goutman, Stephen (Univ of Michigan); Karam, Chafic (Oregon Health Sciences); Lacomis, David (Univ. of Pittsburgh); Manousakis, George (Univ. of Minnesota); Miller, Timothy (Washington Univ St. Louis); Moreno, Cristiane (Columbia University); Pal, Suvankar (Univ. of Edinburgh); Sareen, Dhruv (Cedars Sinai); Simmons, Zachary (Penn State Hershey); Wang, Leo (Univ of Washington). Funding was provided by the ALS Association (National); ALS Association (Greater New York Chapter), and Biogen.

We would like to acknowledge ALS patients and their families. Samples and associated phenotype data used in this study were provided by the ALS Consortium members such as but not limited to University of California San Diego, Johns Hopkins University, Columbia University Medical Center, Barrow Neurological Institute, University of Edinburgh, Georgetown University, University College London, Massachusetts General Hospital, Academic Medical Center, Mount Sinai, Tel-Aviv Sourasky Medical Center, University of Pennsylvania, Penn State University, Washington University, Henry Ford Health Systems, Cedars- Sinai Medical Center, University of Thessaly, University of Athens, Hadassah Medical Center. We would like to acknowledge the ALS Association (ALSA, 19-SI-459) and the Tow Foundation for providing financial support to carry out the sequencing. We would like to acknowledge The Target ALS Human Postmortem Tissue Core https://www.targetals.org/ for providing human postmortem tissue.

---

**Figure S1.**
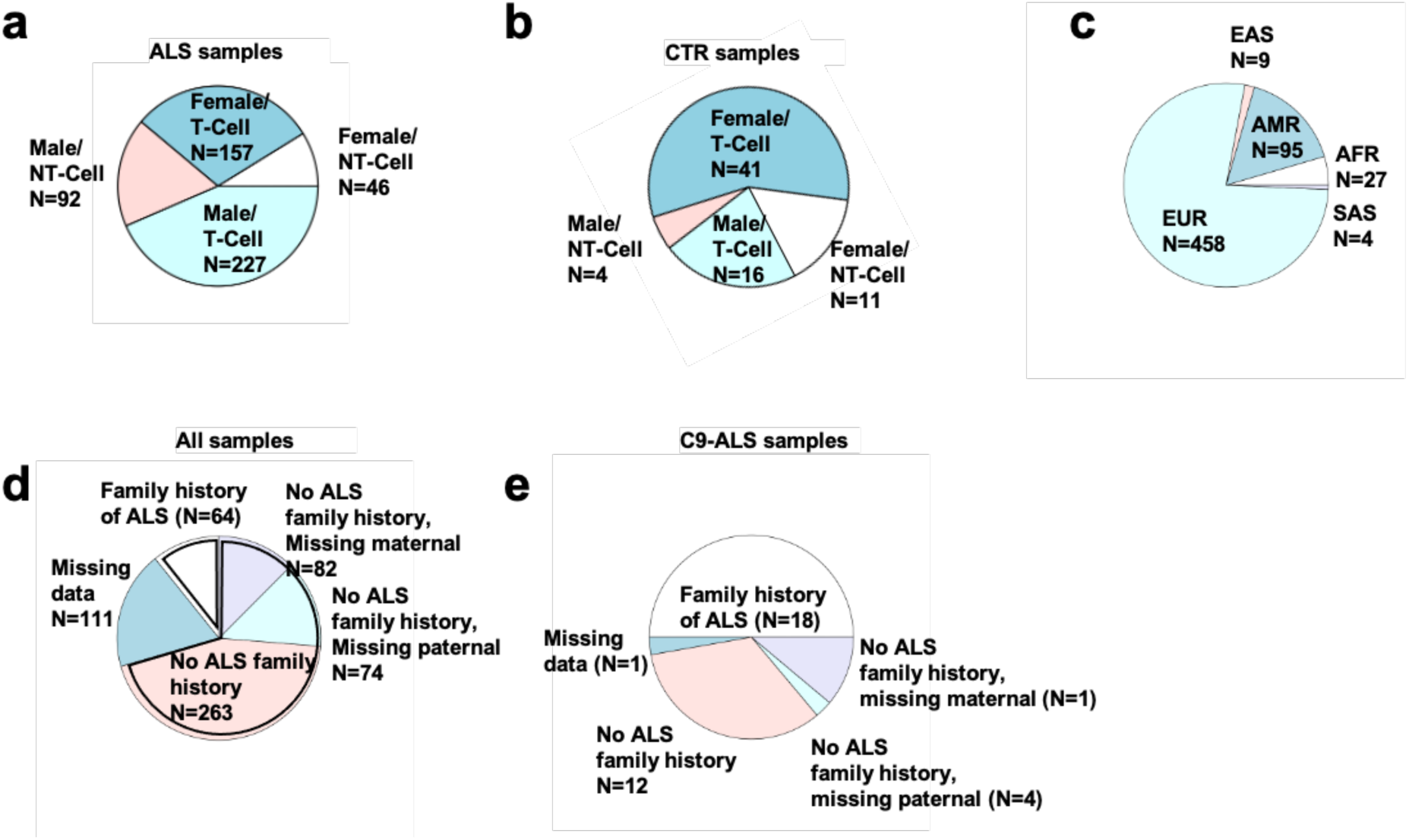
Demographics of AALS subjects used in QTL analysis. A) Pie chart depicting demographics of ALS samples by sex and iPSC cell type of origin (T-cell/NT-cell derived). B) Same as (A), but for healthy control lines. C) Proportions by ancestry. EUR: European ancestry, EAS: East Asian ancestry, AMR: Native American ancestry, AFR: African ancestry, SAS: Southeast Asian ancestry. D) Distribution of data availability on family history for all ALS and control samples. “No ALS family history” means that data for at least one maternal and one paternal relative is present. E) Distribution of family history availability for all subjects carrying the C9orf72 mutant hexanucleotide repeat expansion.

**Figure S2.**
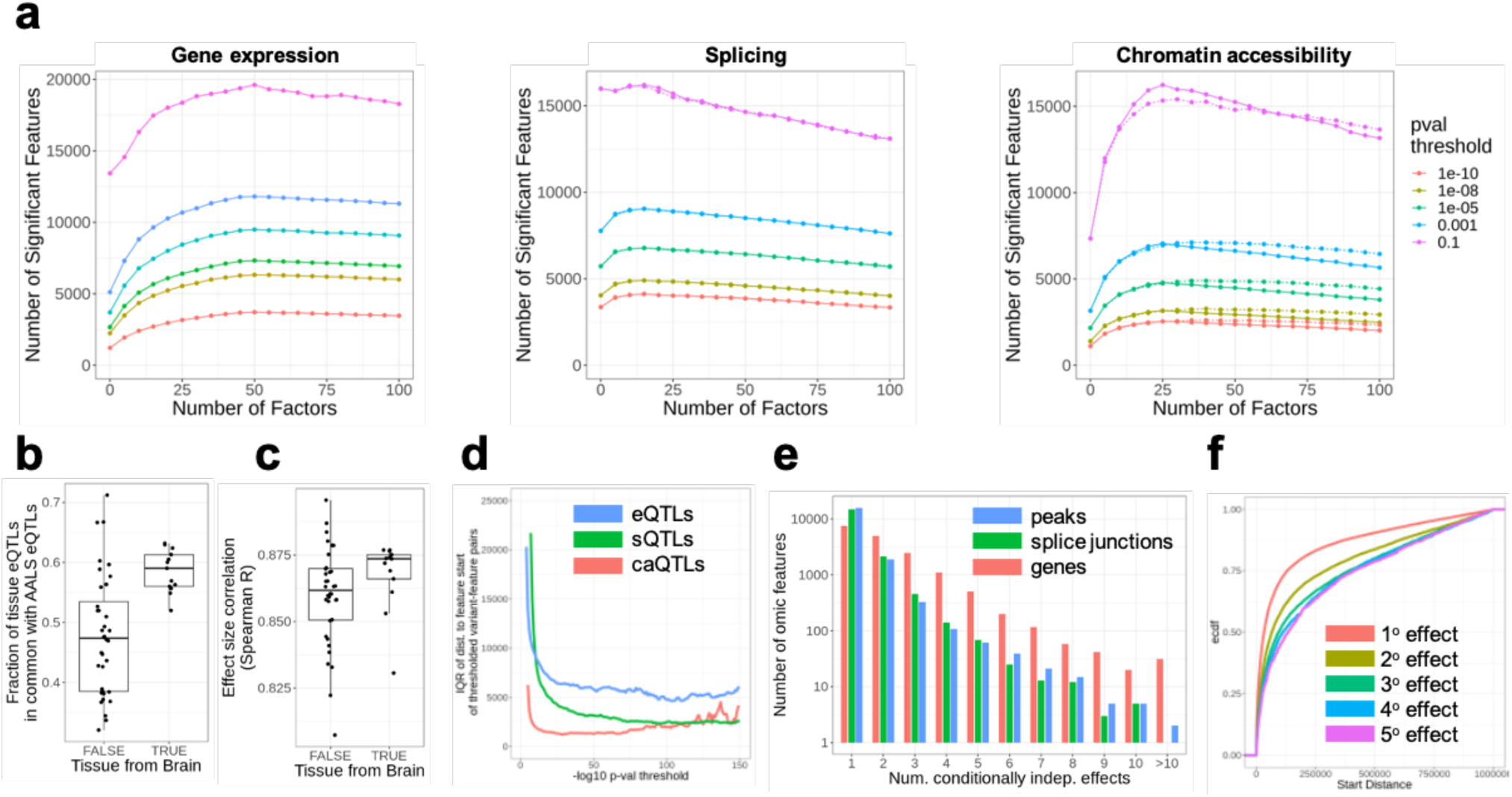
Extended QTL characterization and accounting for batch effects. A) Plots showing the number of significant features (y axis) as a function of the number of top PEER factors regressed out (x axis) for eQTLs (left), sQTLs (middle), and caQTLs (right). Colored lines represent p-value thresholds (see inserted legend). B) Fraction overlap between GTEx Consortium tissue-specific eQTLs and AALS eQTLs, for 47 tissues. C) Effect size correlation between GTEx Consortium tissue-specific eQTL and AALS eQTLs, for 47 tissues. D) Interquartile range of the distance to feature for eQTLs, sQTLs, and caQTLs as a function of p-value threshold. Colored lines represent various QTL (see inserted legend). E) Distribution of the number of conditionally independent effects (genomic variants) influencing gene expression, splicing, and chromatin accessibility. Colored lines represent various effectors (see inserted legend). F) Empirical cumulative distribution function of the distance to the transcription start site for primary eQTLs (1° effect, top driver of variation), secondary eQTLs (2° effect, second driver of variation), etc. (see methods). Colored lines represent various effects (see inserted legend).

**Figure S3:**
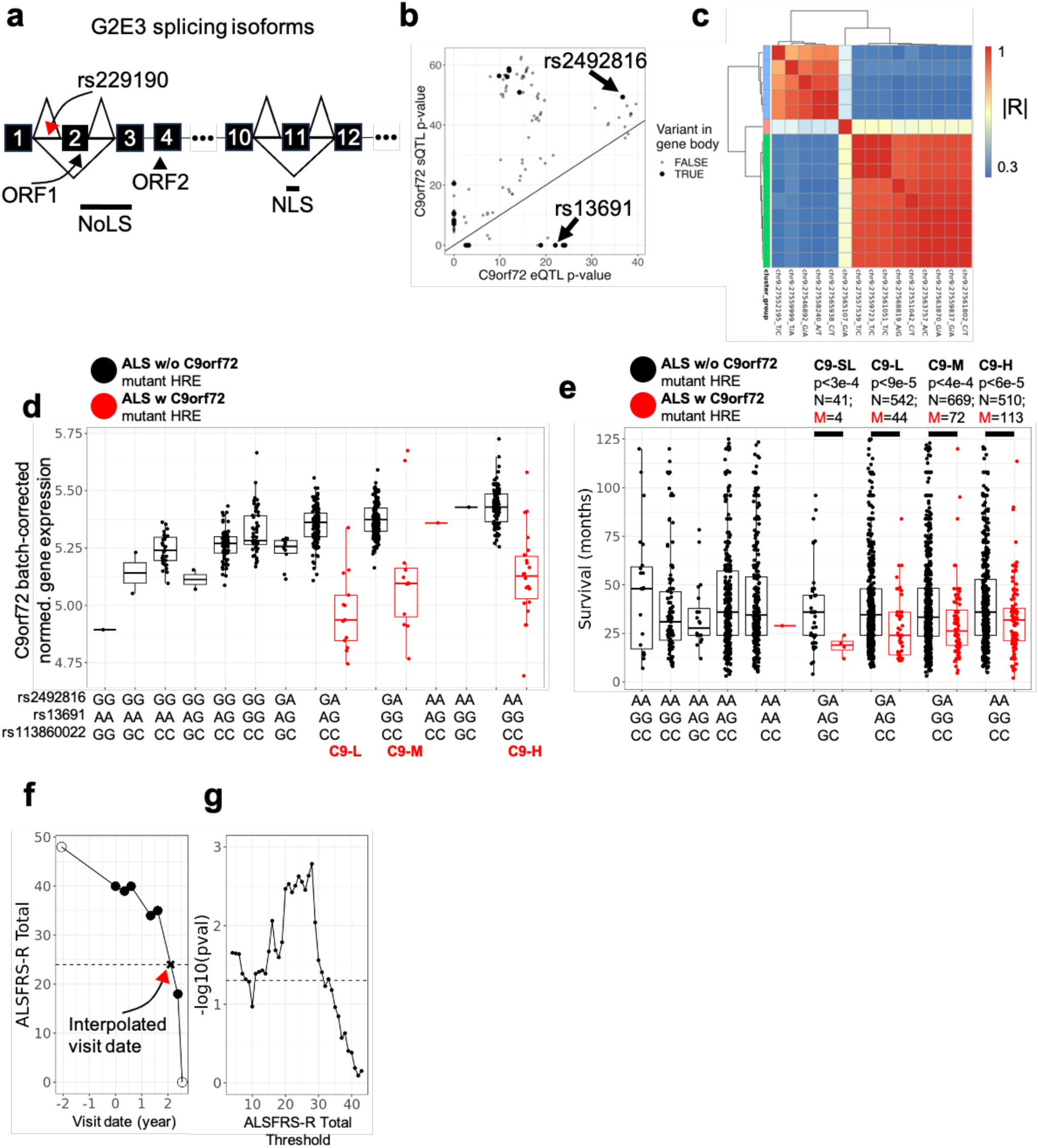
Comparison of significant QTLs and significant ALS GWAS variants. A) Schematic illustrating alternative splicing of *G2E3*. ORF: open reading frame. NoLS: Nucleolar localization sequence. NLS: nuclear localization sequence. Skipping of exon 2 is significantly associated with rs229190 genotype. There is frequent skipping of exon 11, but no significant associations with genotype could be found. B) *C9orf72* sQTL p-values plotted against *C9orf72* eQTL p-values. Large solid dots correspond to QTLs in the pre-mRNA transcript; small unfilled dots are not. C) Correlation matrix of genomic variants from Figure 3f that fell within the gene body and were either highly significant sQTLs or eQTLs. Variants in cluster group 1 were only highly significant sQTLs, variants in cluster group 2 were only highly significant eQTLs, and the variant in cluster group 3 was both a highly significant eQTL and sQTL. D) Batch-corrected normalized gene expression of *C9orf72* plotted as a function of the combined genotypes of rs2492816 (top genotype), rs13691 (middle genotype), and rs113860022 (bottom genotype). Red dots indicate carriers of the mutant repeat expansion; black dots indicate non-C9 ALS patients and healthy controls. C9-L, C9-M, and C9-H label annotate the C9-ALS patients by the haplotype group of their wild-type allele (see Figure 4E). E) Disease survival plotted as a function of the combined genotypes of rs2492816 (top genotype), rs13691 (middle genotype), and rs113860022 (bottom genotype) for all ALS patients with available data in the ALS Compute database. Genotypes are plotted to match Figure 4E to the extent possible. F) Example estimation of time to ALSFRS-R = 24. Solid points indicate gathered longitudinal data. Empty point indicates date of death, anchored at 0. Dashed horizontal line corresponds to ALSFRS-R=24. G) The p-value as calculated for Figure 4j, but with a range of different ALSFRS-R total thresholds. Dashed line corresponds to p-value 0.05.

**Figure S4.**
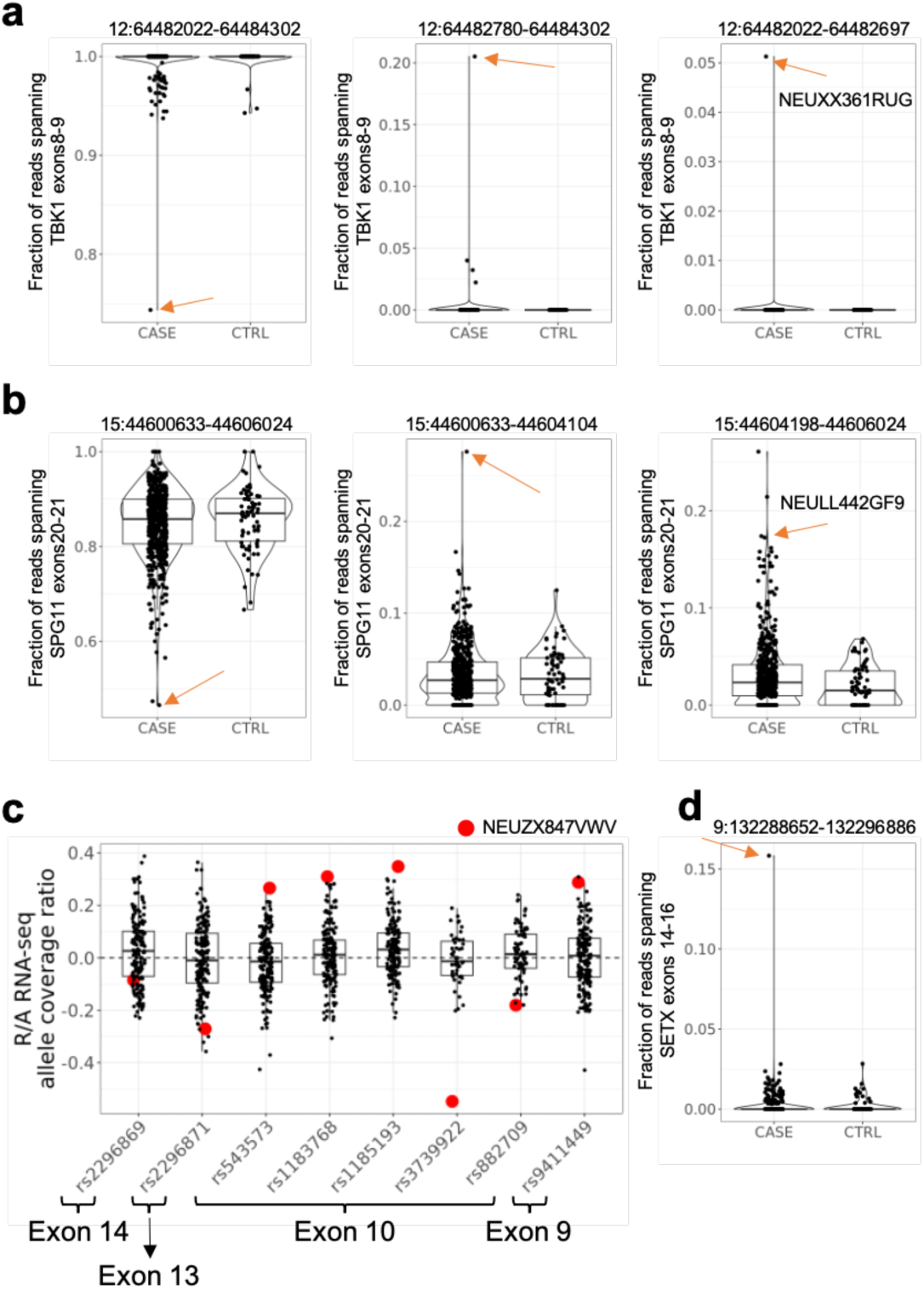
Rare splicing in ALS genes. A) Percent spliced in for cryptic exon in *TBK1* found in sample NEUXX361RUG. Red arrow indicates NEUXX361RUG. B) Percent spliced in for cryptic exon found in *SPG11* in sample NEULL442GF9. Red arrow indicates NEULL442GF9. C) RNA-seq reference/alternate allele imbalance across all samples for eight SNPs in SETX at which NEUZX847VWV (red) is heterozygous. D) Percent spliced in for *SETX* exon 15. Red arrow indicates NEUZX847VWV.

## Supplementary Table Captions

Table S1:

Metadata for 594 iPSC-derived motor neuron lines used in this study.

Table S2:

Table of top eQTLs for each gene, identified using a permutation test.

Table S3:

Table of top sQTLs for each intron cluster, identified using a permutation test.

Table S4:

Table of top caQTLs for each chromatin peak, identified using a permutation test.

Table S5:

Table of top conditionally independent eQTLs for each gene.

Table S6:

Table of top conditionally independent sQTLs for each intron cluster.

Table S7:

Table of top conditionally independent caQTLs for each chromatin peak.

Table S8:

Table of significant eQTLs that are also Project MinE GWAS variants.

Table S9:

Table of significant sQTLs that are also Project MinE GWAS variants.

Table S10:

Table of significant caQTLs that are also Project MinE GWAS variants.

Table S11:

Table of significant C9orf72 eQTLs identified after rerunning QTL analysis on subset of samples that do not include mutant repeat expansion carriers.

Table S12:

Table of significant C9orf72 exon 1a/1b sQTLs identified after rerunning QTL analysis on subset of samples that do not include mutant repeat expansion carriers.

Table S13:

Table of significant C9orf72 promoter chromatin peak caQTLs identified after rerunning QTL analysis on subset of samples that do not include mutant repeat expansion carriers.

Table S14:

List of ALS Compute sample IDs with available survival data at month-resolution used for C9-ALS subgroup analysis.

Table S15:

List of ALS Compute and Answer ALS sample IDs used for longitudinal ALSFRS-R Total C9-ALS subgroup analysis.

Table S16:

List of differential accessibly genomic positions significantly associated with the genotype of rs2492816 from ATAC-seq data at single base pair resolution.

## References

1. van Es, M. A. et al. Amyotrophic lateral sclerosis. The Lancet 390, 2084–2098 (2017).

2. Goutman, S. A. et al. Emerging insights into the complex genetics and pathophysiology of amyotrophic lateral sclerosis. The Lancet Neurology 21, 465–479 (2022).

3. Chia, R., Chiò, A. & Traynor, B. J. Novel genes associated with amyotrophic lateral sclerosis: diagnostic and clinical implications. The Lancet Neurology 17, 94–102 (2018).

4. Van Rheenen, W. et al. Common and rare variant association analyses in amyotrophic lateral sclerosis identify 15 risk loci with distinct genetic architectures and neuron-specific biology. Nat Genet 53, 1636–1648 (2021).

5. Brown, A.-L. et al. TDP-43 loss and ALS-risk SNPs drive mis-splicing and depletion of UNC13A. Nature 603, 131–137 (2022).

6. Baxi, E. G. et al. Answer ALS, a large-scale resource for sporadic and familial ALS combining clinical and multi-omics data from induced pluripotent cell lines. Nature Neuroscience 25, 226–237 (2022).

7. Mizielinska, S. et al. Amyotrophic lateral sclerosis caused by hexanucleotide repeat expansions in C9orf72: from genetics to therapeutics. The Lancet Neurology 24, 261–274 (2025).

8. Tsitkov, S. et al. Disease related changes in ATAC-seq of iPSC-derived motor neuron lines from ALS patients and controls. Nat Commun 15, 3606 (2024).

9. Workman, M. J. et al. Large-scale differentiation of iPSC-derived motor neurons from ALS and control subjects. Neuron 111, 1191–1204.e5 (2023).

10. ALS Compute. dbGaP.

11. Genomic Translation for ALS Care (GTAC) - WGS. dbGaP.

12. DEMENTIA-SEQ: WGS in Lewy Body Dementia and Frontotemporal Dementia. dbGaP.

13. Using NGS to Sequence Whole Genomes to Identify Genes Underlying ALS. dbGaP.

14. Identification of ALS Associated Genes Using Whole Genome Sequencing. dbGaP.

15. Stegle, O., Parts, L., Piipari, M., Winn, J. & Durbin, R. Using probabilistic estimation of expression residuals (PEER) to obtain increased power and interpretability of gene expression analyses. Nat Protoc 7, 500–507 (2012).

16. Dolzhenko, E. et al. ExpansionHunter: a sequence-graph-based tool to analyze variation in short tandem repeat regions. Bioinformatics 35, 4754–4756 (2019).

17. Wang, D. et al. Comprehensive functional genomic resource and integrative model for the human brain. Science 362, eaat8464 (2018).

18. The GTEx Consortium et al. The GTEx Consortium atlas of genetic regulatory effects across human tissues. Science 369, 1318–1330 (2020).

19. Saez-Atienzar, S. et al. Genetic analysis of amyotrophic lateral sclerosis identifies contributing pathways and cell types. Sci. Adv. 7, eabd9036 (2021).

20. Brooks, W. S., Banerjee, S. & Crawford, D. F. G2E3 is a nucleo-cytoplasmic shuttling protein with DNA damage responsive localization. Experimental Cell Research 313, 665–676 (2007).

21. Grandi, F. C., Modi, H., Kampman, L. & Corces, M. R. Chromatin accessibility profiling by ATAC-seq. Nat Protoc 17, 1518–1552 (2022).

22. Rauluseviciute, I. et al. JASPAR 2024: 20th anniversary of the open-access database of transcription factor binding profiles. Nucleic Acids Research 52, D174–D182 (2024).

23. Murphy, N. A. et al. Age-related penetrance of the C9orf72 repeat expansion. Sci Rep 7, 2116 (2017).

24. Smith, B. N. et al. The C9ORF72 expansion mutation is a common cause of ALS+/−FTD in Europe and has a single founder. Eur J Hum Genet 21, 102–108 (2013).

25. Majounie, E. et al. Frequency of the C9orf72 hexanucleotide repeat expansion in patients with amyotrophic lateral sclerosis and frontotemporal dementia: a cross-sectional study. The Lancet Neurology 11, 323–330 (2012).

26. Cooper-Knock, J. et al. Rare Variant Burden Analysis within Enhancers Identifies CAV1 as an ALS Risk Gene. Cell Reports 33, 108456 (2020).

27. Al Khleifat, A. et al. Structural variation analysis of 6,500 whole genome sequences in amyotrophic lateral sclerosis. npj Genom. Med. 7, 8 (2022).

28. Megat, S. et al. CREB3 gain of function variants protect against ALS. Nat Commun 16, 2942 (2025).

29. Cady, J. et al. Amyotrophic lateral sclerosis onset is influenced by the burden of rare variants in known amyotrophic lateral sclerosis genes. Annals of Neurology 77, 100–113 (2015).

30. Landrum, M. J. et al. ClinVar: public archive of interpretations of clinically relevant variants. Nucleic Acids Res 44, D862–D868 (2016).

31. Van Daele, S. H. et al. Genetic variability in sporadic amyotrophic lateral sclerosis. Brain 146, 3760–3769 (2023).

32. Sherry, S. T. dbSNP: the NCBI database of genetic variation. Nucleic Acids Research 29, 308–311 (2001).

33. Van Blitterswijk, M. et al. Association between repeat sizes and clinical and pathological characteristics in carriers of C9ORF72 repeat expansions (Xpansize-72): a cross-sectional cohort study. The Lancet Neurology 12, 978–988 (2013).

34. Jackson, J. L. et al. Elevated methylation levels, reduced expression levels, and frequent contractions in a clinical cohort of C9orf72 expansion carriers. Mol Neurodegeneration 15, 7 (2020).

35. Fournier, C. et al. Relations between C9orf72 expansion size in blood, age at onset, age at collection and transmission across generations in patients and presymptomatic carriers. Neurobiology of Aging 74, 234.e1–234.e8 (2019).

36. Van Blitterswijk, M. et al. TMEM106B protects C9ORF72 expansion carriers against frontotemporal dementia. Acta Neuropathol 127, 397–406 (2014).

37. on behalf of the BELNEU CONSORTIUM et al. The C9orf72 repeat size correlates with onset age of disease, DNA methylation and transcriptional downregulation of the promoter. Mol Psychiatry 21, 1112–1124 (2016).

38. Waite, A. J. et al. Reduced C9orf72 protein levels in frontal cortex of amyotrophic lateral sclerosis and frontotemporal degeneration brain with the C9ORF72 hexanucleotide repeat expansion. Neurobiology of Aging 35, 1779.e5–1779.e13 (2014).

39. Shi, Y. et al. Haploinsufficiency leads to neurodegeneration in C9ORF72 ALS/FTD human induced motor neurons. Nat Med 24, 313–325 (2018).

40. DeJesus-Hernandez, M. et al. Expanded GGGGCC Hexanucleotide Repeat in Noncoding Region of C9ORF72 Causes Chromosome 9p-Linked FTD and ALS. Neuron 72, 245–256 (2011).

41. Zhu, Q. et al. Reduced C9ORF72 function exacerbates gain of toxicity from ALS/FTD-causing repeat expansion in C9orf72. Nat Neurosci 23, 615–624 (2020).

42. Boivin, M. et al. Reduced autophagy upon C9ORF72 loss synergizes with dipeptide repeat protein toxicity in G4C2 repeat expansion disorders. The EMBO Journal 39, e100574 (2020).

43. Liu, Y. et al. DNA-initiated epigenetic cascades driven by C9orf72 hexanucleotide repeat. Neuron 111, 1205–1221.e9 (2023).

44. Lall, D. et al. C9orf72 deficiency promotes microglial-mediated synaptic loss in aging and amyloid accumulation. Neuron 109, 2275–2291.e8 (2021).

45. Van Den Berg, L. H., et al. Safety, tolerability, and pharmacokinetics of antisense oligonucleotide BIIB078 in adults with C9orf72-associated amyotrophic lateral sclerosis: a phase 1, randomised, double blinded, placebo-controlled, multiple ascending dose study. The Lancet Neurology 23, 901–912 (2024).

46. Chew, J., et al. *C9ORF72* repeat expansions in mice cause TDP-43 pathology, neuronal loss, and behavioral deficits. Science 348, 1151–1154 (2015).

47. Elia, L. P., Mason, A. R., Alijagic, A. & Finkbeiner, S. Genetic Regulation of Neuronal Progranulin Reveals a Critical Role for the Autophagy-Lysosome Pathway. J. Neurosci. 39, 3332–3344 (2019).

48. Hashimoto, K., Jahan, N., Miller, Z. A. & Huang, E. J. Neuroimmune dysfunction in frontotemporal dementia: Insights from progranulin and C9orf72 deficiency. Current Opinion in Neurobiology 76, 102599 (2022).

49. Freischmidt, A. et al. Haploinsufficiency of TBK1 causes familial ALS and fronto-temporal dementia. Nat Neurosci 18, 631–636 (2015).

50. Rifai, O. M. et al. Clinicopathological analysis of *NEK1* variants in amyotrophic lateral sclerosis. Brain Pathology 35, e13287 (2025).

51. Mann, J. R. et al. Loss of function of the ALS-associated NEK1 kinase disrupts microtubule homeostasis and nuclear import. Sci. Adv. 9, eadi5548 (2023).

52. Maruyama, H. et al. Mutations of optineurin in amyotrophic lateral sclerosis. Nature 465, 223–226 (2010).

53. Kim, G., Gautier, O., Tassoni-Tsuchida, E., Ma, X. R. & Gitler, A. D. ALS Genetics: Gains, Losses, and Implications for Future Therapies. Neuron 108, 822–842 (2020).

54. Mai, X. et al. Structural basis for membrane remodeling by the AP5–SPG11–SPG15 complex. Nat Struct Mol Biol 32, 1334–1346 (2025).

55. Fujii, T. et al. Parkinson’s disease-associated ATP13A2/PARK9 functions as a lysosomal H+,K+-ATPase. Nat Commun 14, 2174 (2023).

56. Spataro, R. et al. Mutations in ATP13A2 (PARK9) are associated with an amyotrophic lateral sclerosis-like phenotype, implicating this locus in further phenotypic expansion. Hum Genomics 13, 19 (2019).

57. Daoud, H. et al. Exome sequencing reveals SPG11 mutations causing juvenile ALS. Neurobiology of Aging 33, 839.e5–839.e9 (2012).

58. Park, J., Blair, N. F. & Sue, C. M. The role of ATP13A2 in Parkinson’s disease: Clinical phenotypes and molecular mechanisms. Movement Disorders 30, 770–779 (2015).

59. Richard, P. et al. SETX (senataxin), the helicase mutated in AOA2 and ALS4, functions in autophagy regulation. Autophagy 17, 1889–1906 (2021).

60. Nanetti, L. et al. SETX mutations are a frequent genetic cause of juvenile and adult onset cerebellar ataxia with neuropathy and elevated serum alpha-fetoprotein. Orphanet J Rare Dis 8, 123 (2013).

61. Hadjinicolaou, A. et al. De novo pathogenic variant in SETX causes a rapidly progressive neurodegenerative disorder of early childhood-onset with severe axonal polyneuropathy. acta neuropathol commun 9, 194 (2021).

62. Shiloh, Y. The ATM-mediated DNA-damage response: taking shape. Trends in Biochemical Sciences 31, 402–410 (2006).

63. Ramachandran, S. et al. Hypoxia-induced SETX links replication stress with the unfolded protein response.Nat Commun 12, 3686 (2021).

64. Taylor-Weiner, A. et al. Scaling computational genomics to millions of individuals with GPUs. Genome Biol 20, 228 (2019).

65. Kumar, S., Ambrosini, G. & Bucher, P. SNP2TFBS – a database of regulatory SNPs affecting predicted transcription factor binding site affinity. Nucleic Acids Res 45, D139–D144 (2017).

66. Martin Morgan, H. P. Rsamtools. Bioconductor 10.18129/B9.BIOC.RSAMTOOLS (2017).

67. Wang, G., Sarkar, A., Carbonetto, P. & Stephens, M. A Simple New Approach to Variable Selection in Regression, with Application to Genetic Fine Mapping. Journal of the Royal Statistical Society Series B: Statistical Methodology 82, 1273–1300 (2020).

68. Love, M. I., Huber, W. & Anders, S. Moderated estimation of fold change and dispersion for RNA-seq data with DESeq2. Genome Biol 15, 550 (2014).

69. Rausch, T. et al. DELLY: structural variant discovery by integrated paired-end and split-read analysis. Bioinformatics 28, i333–i339 (2012).

